# Molecular Profiling of Breast Cancer Susceptibility of Obese or Insulin-Resistant and Pre-Diabetic Patients Using ITLN1 and CD295 SNPs

**DOI:** 10.1101/2020.01.08.20016980

**Authors:** Nadia M. Hamdy, Reham A. El-Shemy

## Abstract

Mutations in cluster of differentiation (CD) 295 gene, encoding class I cytokine receptor, are associated with obesity and breast cancer (BC). SNPs in the adipocyte-inferred novel cytokine intelectin 1 (ITLN1) remain understudied in connection to CD295 polymorphisms and diabetes mellitus (DM) or a pre-diabetic state, as well as to DNA damage seen in BC. We will explore whether CD295 (ID rs6700896) and ITLN1 (rs rs952804) SNPs impact BC with or without DM, insulin resistance (IR) or obesity. Effects of ITLN1 or CD295 polymorphism(s) on DNA damage in BC were also examined. Blood samples from 170 women with BC (including 33 and 48 with DM and pre-diabetes, respectively) and from 108 age-matched women in the control group were collected. Plasma insulin, leptin, CD295, and ITLN1 levels were measured by ELISA. DNA damage was assessed using an alkaline comet assay.

BC cases with clinical stage T II and positive LN as well as tumor histologic grade III, presence of obesity, pre-diabetic events, DM or IR were associated with CD295 rs6700986 mutant homozygous (CC) and heterozygous (CT) genotype and ITLN1 rs952804 mutant heterozygous genotype (CT) (P ≤ 0.05). Tail DNA (%) and tail moment units were significantly associated with CD295 rs6700986 CT and ITLN1 rs952804 TT genotypes. C allele (CT+CC vs. TT) and T allele (TT+CT vs. CC) for CD295 rs6700986 and ITLN1 rs952804, respectively, were associated with BC risk (P ≤ 0.05). ITLN1 (rs952804) and CD295 (rs6700986) SNPs should be considered as BC associated-susceptibility risk factors in obese, insulin resistant, or pre-diabetics.

**Graphical Abstract:** 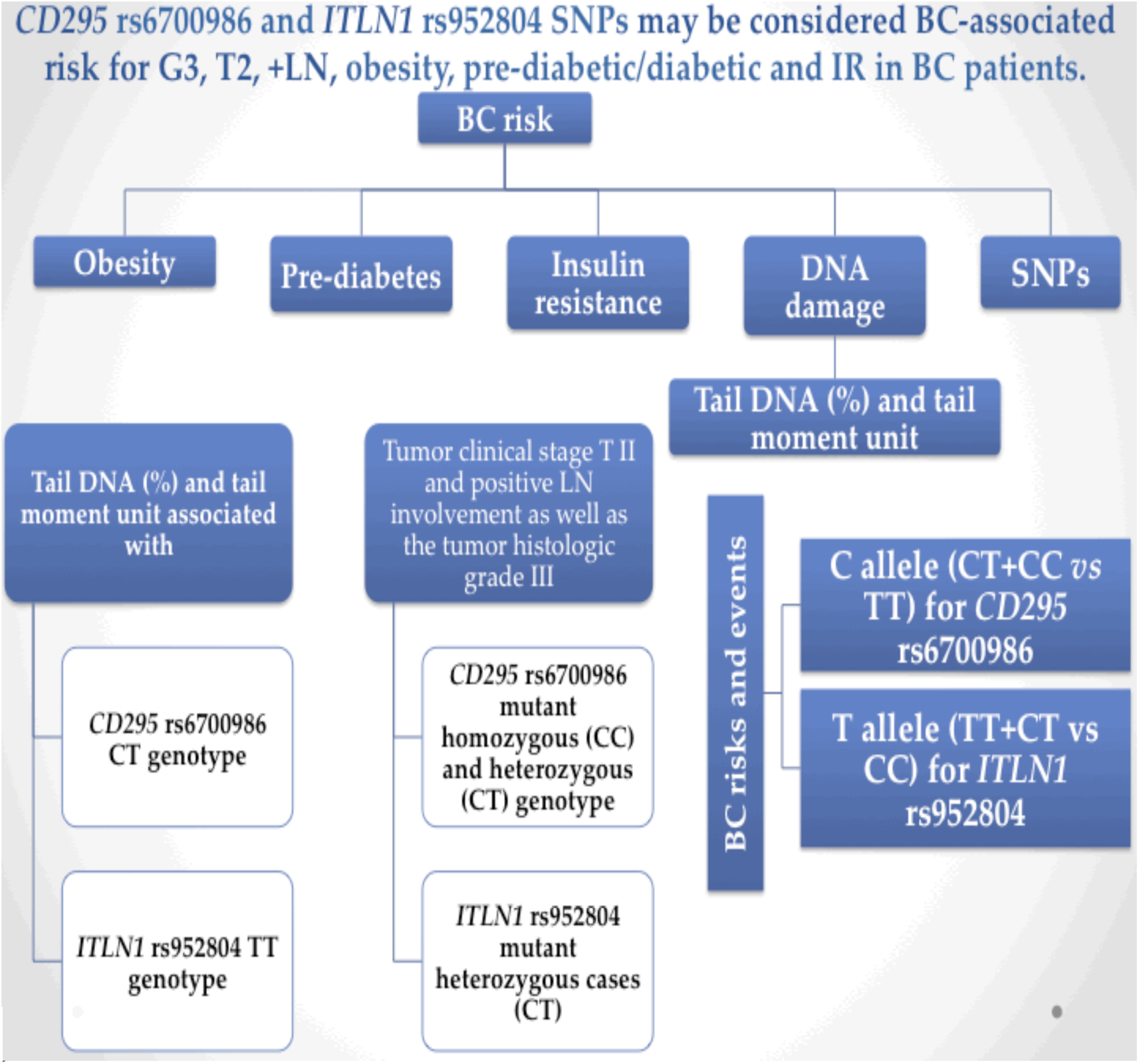

**Remarks/Highlights:**

- *ITLN1* and *CD295* polymorphism testing might be utilized for accessing BC susceptibility in either obese or insulin resistant, pre-diabetic patients.
- A modestly increased risk of BC in women harboring the C allele of *CD295* rs6700986 polymorphism and the T allele of the *ITLN1* rs952804, where:

1. Tumor clinical stage T II and positive LN involvement as well as the tumor histologic grade III, obesity existence, pre-diabetic event and being diabetic as well as IR cases were associated with *CD295* rs6700986 mutant homozygous (CC) and heterozygous (CT) genotype and *ITLN1* rs952804 mutant heterozygous cases (CT) (*P* ≤ 0.05).
2. Tail DNA (%) and tail moment unit were significantly associated with *CD295* rs6700986 CT genotype and *ITLN1* rs952804 TT. These SNPs could be considered as BC associated risk factor.

- In dominant effect of the C allele (CT+CC *vs* TT) and the T allele (TT+CT vs CC) for *CD295* rs6700986 and *ITLN1* rs952804, respectively, were associated with BC events and risk (*P* ≤ 0.05).
- *CD295* rs6700986 and *ITLN1* rs952804 SNPs may be considered BC-associated risk for G3, T2, +LN, obesity, pre-diabetic/diabetic and IR in BC patients.

## 1. BACKGROUND

Breast cancer (BC) is a very common cancer (Ibrahim et al 2014, Adeloye et al 2018). BC cases that have high levels of immune cells and/or elevated levels of pro-inflammatory cytokines are anticipated to have a poor prognosis (Stender et al 2017). Adipokines are proteins secreted by adipose tissue that are involved in a wide range of processes, including insulin sensitivity (Miehle et al 2012). A large body of evidence has supported the role of adipose tissue in the regulation of insulin resistance (IR). Meanwhile, adipocytokines, which are adipocyte-derived hormones, have also been implicated in the regulation of metabolism and IR. The protein encoded by the cluster of differentiation *(CD)295* gene belongs to a family of cytokine receptors. There are more than 50 cytokines in the adipokine family, including CD295, a class I cytokine receptor, which play roles in malignancy in obese individuals (Vona-Davis and Rose 2007). Long chain CD295 receptor type b is responsible for most of the effects mediated by CD295 (Laschober et al 2009). CD295 is known to stimulate gene transcription by activating the cytosolic JAK-STAT proteins MAPK, PI3K, and AMPK (Mahmoudi et al 2015). CD295 is also a receptor for leptin, an adipocyte-specific hormone that regulates body weight, and is involved in regulating metabolism in adipose tissues, as well as in a novel hematopoietic pathway that is required for normal lymphopoiesis. The adipocyte-inferred cytokine intelectin 1 (ITLN1) is a novel secretory and galactose-binding lectin that appears to have an anti-inflammatory role in both obesity (Jamshidi et al 2017) and inflammation-related diseases (Huang et al 2016), including BC. The *ITLN1* gene product omentin1 is mainly expressed in visceral adipose tissue, and was shown to inhibit inflammatory responses and mitigate IR as well as other obesity-related disorders (Du et al 2016). An immediate relationship between obesity and increased risk of BC has been reported (Imayama et al 2013). The relationship of BC to inflammation and obesity suggests that polymorphisms in the CD295 and/or *ITLN1* genes, which both play roles in adipose tissues, could occur BC. Moreover, hyperinsulinemia actuates proliferative tissue anomalies, due to the strong anabolic impact of insulin, which stimulates DNA synthesis and cell proliferation (Boyd 2003). Increased IR promotes elevated glucose levels that can be specifically cytotoxic and pro-inflammatory (Nasiri et al., 2019), as well as oxidatively stressful, and thus could be associated with DNA damage (Ibarra-Drendall et al 2011).

This study was conducted to determine whether single nucleotide polymorphisms (SNPs) in *CD295* (ID rs6700896) and *ITLN1* (rs rs952804) confer risk for metabolic disorders (IR and/or DM) in female BC patients in Egypt. The first aim of this study was to explore how these SNPs affect BC in the presence or absence of diabetes, insulin resistance, or obesity, whereas the second aim was to examine DNA damage in these patients using a comet test to detect DNA strand breaks in individual cells from mononuclear cells isolated from blood samples (Galardi et al 2012). Realizing that no study inspected the relationship of polymorphisms in these genes with the level of DNA damage in BC cases, this will be the first main study to do this examination.

## 2. SUBJECTS, MATERIALS, AND METHODS

### 2.1. Subjects

This is a retrospective randomized single-center study, composed of patients treated with chemotherapy for inflammatory BC at a University center (National Cancer Institute (NCI), Cairo) between January 2015 and December 2015. The study protocol was approved by the ethical committees of the Faculty of Pharmacy, Ain Shams University (code number 30/7/2016-F-EBD-01-03) and the NCI, Cairo University,

Cairo, Egypt. The study was carried out in accordance with the regulations and recommendations of the Declaration of Helsinki (WMA 2013). Written informed consent was obtained from all study subjects.

#### 2.1.1. Inclusion criteria

Females diagnosed with invasive ductal carcinoma (IDC) for the first time at age 43 or older were included. A total of 170 postmenopausal female patients aged 43-75 were selected and the median age was 53 years-old.

### 2.1.2. Exclusion criteria

Patients having incomplete data or histopathologic diagnosis, as well as patients with metastasis at the time of initial diagnosis were excluded. Males with BC were excluded as were BC types other than IDC. Other exclusion criteria were: previous or current history of acute or chronic viral hepatitis; malignant disease; acute infections; pituitary, adrenal, thyroid, or pancreatic disease, or evidence of any other endocrine disorders; or prolonged use of corticosteroids or sex hormones.

#### 2.1.3. Control group

A group of 108 age-matched healthy controls (median age 53 years) with normal liver enzyme levels and no clinical or laboratory evidence of BC was recruited during routine wellness examinations.

#### 2.1.4. Participant data

The demographic characteristics of the study subjects are summarized in Table 1a. Clinical data were obtained using medical records and original pathology reports and were compiled in an Excel database (Microsoft Corporation, Redmond, WA, USA). The following parameters were assessed: patient age, tumor size (defined as sonographic diameter (mm) on diagnosis), initial tumor stage and nodal status according to TMN classification, histologic subtype, estrogen receptor status, progesterone status, HER2 status (a score of 0 to +1 was regarded as HER2 negative and a score of +3 as positive), grading and proliferation status as assessed by Ki-67 staining, and chemotherapy regimen (doxorubicin/trastuzumab/lapatinib). All histopathological parameters included were derived from the original pathology reports. Scoring was performed by a specialized pathologist at the Pathology Department, NCI, Cairo University, according to standardized protocols.

**Table 1a.** Anthropometric parameters, obesity and diabetes factors, and breast cancer clinical factors in breast cancer patients compared to the control group.

| Parameter | Control (n=108) | BC patients (n=170) | *P-value* |
| --- | --- | --- | --- |
| Age (years) | 53 (43-66) | 53 (43-75) |  |
| Insulin (μIU/mL) | 8 (4.5-19) | 10.6 (3.8-17.3) | 0.007 |
| FBG (mg/dL) | 100 (66-144) | 120 (85-224) |  |
| HOMA-IR index | 2.2 (0.81-4.9) | 3.4 (0.9-6.6) | 0.02 |
| HOMA-B | 0.8 (0.31-6) | 0.68 (0.13-1.94) | 0.03 |
| Obese (-/+) | 40/68 | 38/132 |  |
| IR (-/+) | 50/58 | 12/158 |  |
| Diabetes (-/pre/+) | 90/16/2 | 88/48/33 |  |
| Leptin (ng/mL) | 7 (2-11) | 15 (8-50) | <0.001 |
| LepR (ng/mL) | 22 (11-27) | 20 (13-27) | 0.001 |
| ITLN1 (ng/mL) | 99 (66-177) | 40 (30-66) | <0.001 |
| Ejection fraction % |  | 64 (54-80) |  |
| CEA (ng/mL) |  | 4 (1-10) |  |
| CA15.3 (U/mL) |  | 20 (1-126) |  |
| Ki-67 |  | 70 (15-80) |  |
| LH (IU/L) <sup>a</sup> | | 9.9 ± 0.12 | |
| FSH (mIU/L) <sup>a</sup> | | 24.5 ± 1.85 | |
| Estradiol (pg/mL) <sup>a</sup> | | 27.1 ± 6.9 | |
| Tumor clinical stage (T) (I/II) |  | 69/101 |  |
| Lymph node status (N) (-/+) |  | 66/104 |  |
| Metastasis (M) (-/+) |  | 170/0 |  |
| Tumor histological grades (II/III) |  | 141/29 |  |
| BIRAD (III/IV) |  | 42/128 |  |

Data are median (range), statistical analysis was performed using Mann–Whitney U test,
<sup>a</sup> data are mean ± S.D, statistical analysis was performed using Chi-square test.
FBG; fasting blood glucose, HOMA-IR; homeostasis model assessment-insulin resistance, CEA; carcinoembryonic antigen, CA 15.3; cancer antigen 15.3, LH; luteinizing hormone, FSH; follicle stimulating hormone.

### 2.2. Blood sample collection and preparation

A 7 mL blood sample was collected from each patient into 10 mL K3EDTA vacutainer tubes at the end of the clinical examination interview. Samples were stored at -80 °C until biochemical assessment at the Research Lab in the Biochemistry Department, Faculty of Pharmacy, Ain Shams University. One part of the sample was used for DNA extraction from the blood and was carried out using the QIAamp DNA Mini Kit protocol (QIAGEN, Inc., Santa Clarita, CA, USA). The extracted DNA was stored at -80 °C until use in SNP assays. The remaining sample was used for in comet assays.

#### 2.2.1. Biochemical analysis

ELISA was used to determine plasma insulin, leptin, CD295, and ITLN1 levels as well as CEA, CA15.3, luteinizing hormone (LH), and follicle stimulating hormone (FSH) according to the manufacturers’ instructions. Homeostasis model assessment (HOMA), a measure of IR, was calculated as: [(FBG x fasting insulin)/22.5] (Mathews et al 1985). This value is regarded as a simple, inexpensive, and reliable surrogate measure of IR, whereas the HOMA of β-cell function (HOMA-B) index, computed as the product of 20 and basal insulin levels divided by the value of basal glucose concentrations minus 3.5, is accepted as a good measure of β-cell function (Wallace et al 2004).

### 2.3. Genetic analysis

Genotyping was performed using Assays-by-Design supplied by Applied Biosystems International (ABI) (Applied Biosystems, Foster City, CA). Reactions were performed on a 7500 ABI Sequence Detection System. The *CD295* rs6700986 polymorphism is a T/C (FWD) single-nucleotide variation on human chromosome 1:180452403, p31.3, Ancestral Allele: C. GAGAATCACTTGAAACCTAGGAGGC[T/C]AAGGTTGTGGTGAGCCGAGATCATG, Gene: ACBD6 (GeneView), Functional Consequence: intron variant, Global MAF:T=0.4603/2305. The *ITLN1* rs952804 polymorphism is a C/T (FWD) single-nucleotide variation on human chromosome 1:160874955, q23.3, Ancestral Allele: C. CCACCTGCAGCTTTAGAATTGGGTT[C/T]ATCTGTCTTCTCTATCACTTCTTTA, Global MAF:T=0.4335/2171. SNPs (http://www.ncbi.nlm.nih.gov/snp/?term=) were chosen based on a minor allele frequency that exceeded 5%.

#### 2.3.1. Genotyping and single nucleotide polymorphism (SNP)

SNP genotyping was performed using the TaqMan^®^ method for allele-specific detection (ABI), which involves real-time PCR amplification with fluorescence detection (Heid et al 1996). The experiments were performed using an ABI Prism^®^ Sequence Detection System (ABI) and TaqMan^®^ Universal PCR Master Mix (ABI) together with 20 ng genomic DNA template (10 ng/μl). Each 25 μl reaction volume contained 100 ng genomic DNA, 0.2 mM dNTPs, 20 mM Tris-HCl (pH 8.8), 10 mM KCl, 10 Mm (NH4)2SO4, 2 mM MgSO4, 0.1% Triton X-100, and 1 unit *Taq* polymerase (New England BioLabs). PCR amplification conditions were as follows: an initial denaturation step at 94 °C for 5 min followed by 34 cycles of 30 sec at 94 °C; 45 sec at 60 °C and 45 sec at 72 °C; and a final elongation at 72 °C for 10 min. After PCR amplification, we performed an endpoint plate read using an ABI StepOne Plus Real-Time PCR System.

### 2.4. Comet assay

Alkaline comet assays were performed according to a standard protocol (Singh et al., 1988; Comet assay kit, Trevigen). DNA migration in peripheral blood leukocytes was measured using a computer-based image analysis system (COMET IV software, Perceptive) connected to a camera (Leica DFC340 FX). The supplied software computed all major measurement parameters, including tail length, which describes the distance from the center of the nucleoid mass to the distal tail end, tail intensity (TI): the relative fluorescence intensity of the comet tail that is a measure of the percentage of DNA in the tail, and tail moment unit (TMU): essentially the product of tail length and TI.

### 2.5. Statistical analysis

All statistical analyses were conducted using the SPSS statistical package for Windows Version 22.0 (SPSS Software, Chicago, IL, USA). First, we tested the normal distribution of the data in which the results for continuous variables were expressed as mean ± standard deviation (SD) in addition to the median (range) for nonparametric data. To study the association between variables an odds ratio (OR) was calculated. For categorical variables, a Chi-square test was used to make comparisons between different groups. The best cutoff values for the investigated parameter(s) (FSH) were calculated from ROC curves. Genetic analyses were also conducted using unconditional logistic regression. Initially, we used univariate analyses to examine possible associations between each polymorphism and cancer grades (II and III) or T (I and II) or N (-/+), or DNA damage and associated metabolic factors (e.g., insulin, HOMA-IR, and IR). We also conducted subgroup analyses to examine potential interactions by the levels of the pre-specified factors and the studied SNPs. Based on the findings from our population, we computed the frequencies of each allele using Hardy-Weinberg equilibrium (HWE) and online software. A likelihood ratio test was performed to test significances. Correlations were assessed using the Spearman correlation coefficient (*r*). Two-tailed analyses were performed, and *P* values ≤ 0.05 were considered statistically significant.

## 3. RESULTS

A total of 278 individuals were recruited for this study, including 170 females with BC, of whom 158 were insulin resistant (Table 1a). Among the BC patients, 132 were obese. Of the obese BC patients, 88 were normoglycemic, 48 were pre-diabetic and 33 had DM. The healthy control group had 108 individuals, of which 40 and 68 were lean and obese, respectively (Table 1a).

### 3.1. Clinical and biological analysis of the study population

Various anthropometric parameters, obesity and diabetes factors, and BC clinical measures, as well as plasma levels of leptin, CD295, ITLN1 were collected (Table 1a), as was information concerning the SNPs under study and DNA damage parameters for both the BC patients and the healthy controls (Table 1b). There were significant differences among the measured values between the control and BC group in terms of polymorphism frequency and DNA damage (see below; *P* ≤ 0.05).

**Table 1b.**
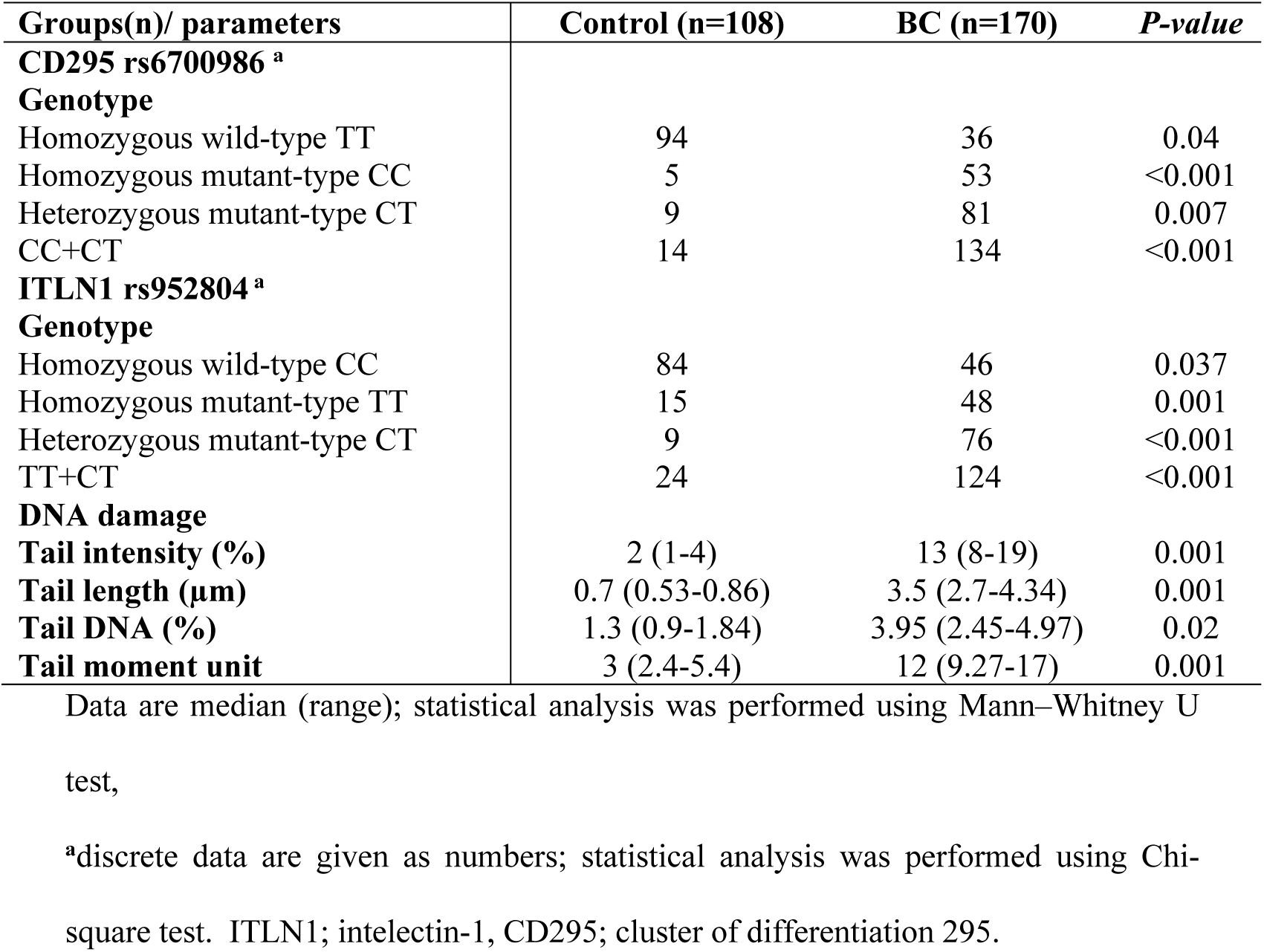
Studied SNPs and DNA damage parameters of breast cancer patients compared to the control group.

### 3.2. Distribution of CD295 and ITLN1 genotypes

*CD295* wild homozygous genotype (TT) was detected in almost all controls (94/108; 87%), but only 36 (21%) BC patients carried this genotype (Table 1b). Meanwhile, mutant homozygous (CC) and heterozygous (CT) *CD295* genotypes were seen in 53 (31.1%) and 81 (47.6) BC cases, respectively. For *ITLN1*, the wild homozygous genotype (CC) was detected in the majority of the control subjects (77.7%), whereas 46 (43%) of BC patients carried this genotype (Table 1b). Mutant homozygous (TT) and heterozygous (CT) genotypes were reported in 48 (28.2%) and 76 (44.7%) BC cases, respectively.

### 3.3. Distribution of the studied SNPs genotypes among the BC patient clinicopathological factors

Stratification of the BC group according to FSH (mIU/L) values revealed that the *CD295* rs6700986 mutant homozygous (CC) and heterozygous (CT) genotype was significantly associated (*P* ≤ 0.05) with BC cases that had FSH values > 25 mIU/L. A similar association was seen for BC patients who were heterozygous for the *ITLN1* rs952804 mutation (CT) (Table 2). HER2/Neu cancer cases with a +3 score had a significantly higher frequency of mutant homozygous (CC) and heterozygous (CT) *CD295* rs6700986 genotypes compared to those who had the wild homozygous (TT) genotype (*P* ≤ 0.05). Similar results were also seen for *ITLN1* rs952804 mutant heterozygous cases (CT).

**Table 2.** Distribution of CD295 SNP rs6700986 and ITLN1 SNP rs952804 genotypes among clinicopathological factors for breast cancer patients (n=170).

| SNP |  | CD295 rs6700986 |  |  |  | ITLN1 rs952804 |  |  |  |
| --- | --- | --- | --- | --- | --- | --- | --- | --- | --- |
|  |  | TT | CC | CT |  | CC | CT | TT |  |
| Clinicopathological factors |  | n (%) | n (%) | n (%) | *P-value\*\*\** | n (%) | n (%) | n (%) | *P-value\*\*\** |
| Breast cancer | Lt | 31 (25.2) | 40 (32.5) | 52 (42.3) | 0.035 | 33 (26.8) | 56 (45.5) | 34 (27.6) |  |
|  | Rt | 5 (10.9) | 12 (26.1) | 29 (63) |  | 13 (28.3) | 19 (41.3) | 14 (30.4) |  |
| FSH (mIU/L) | < 25 | 33 (49.3) | 33 (49) | 68 (51) |  | 18 (39.1) | 23 (30.7) | 19 (39.6) |  |
|  | > 25 | 3 (8.6) | 19 (54.3) | 13 (37) | 0.014 | 28 (60.9) | 52 (69.3) | 29 (60.4) | 0.016 |
| HER2/Neu | 0-1+ | 24 (35.3) | 12 (17.6) | 32 (47.1) |  | 29 (63) | 24 (32) | 15 (31.3) |  |
|  | 2+ | 4 (7.1) | 21 (37.5) | 31 (55.4) |  | 7 (15.2) | 31 (41.3) | 18 (37.5) |  |
|  | 3+ | 8 (17.8) | 19 (42.2) | 18 (40) | 0.001 | 10 (21.7) | 20 (26.7) | 15 (31.3) | 0.004 |
| Tumor clinical stage (T) | I | 13 (19.1) | 24 (35.3) | 31 (45.6) |  | 18 (26.5) | 21 (30.9) | 29 (42.6) |  |
|  | II | 23 (22.8) | 28 (27.7) | 50 (49.5) |  | 28 (27.7) | 54 (53.5) | 19 (18.8) | 0.002 |
| Lymph node status (N) | - | 19 (28.8) | 13 (19.7) | 34 (51.5) |  | 26 (39.4) | 28 (42.4) | 12 (18.2) |  |
|  | + | 17 (16.5) | 39 (37.9) | 47 (45.6) | 0.024 | 20 (19.4) | 47 (45.6) | 36 (35) | 0.007* |
| Tumor histological grade | II | 33 (23.4) | 45 (31.9) | 63 (44.7) |  | 41 (29.1) | 59 (41.8) | 41 (29.1) |  |
|  | III | 3 (10.7) | 7 (25) | 18 (64.3) | 0.032** | 5 (17.9) | 16 (57.1) | 7 (25) | 0.027 |

For FSH the best cutoff value (=25) was calculated by ROC curve.
\* CC different from TT, \*\* CT different from TT, \*\*\* CT different from CC
ITLN1; intelectin-1, FSH; follicle stimulating hormone, CD295; cluster of differentiation 295.

Tumor clinical stage T II and positive LN involvement as well as tumor histologic grade III were associated with *CD295* rs6700986 mutant homozygous (CC) and heterozygous (CT) genotypes, as well as *ITLN1* rs952804 mutant heterozygous cases (CT) (*P* ≤ 0.05; Table 2).

### 3.4. Distribution of CD295 rs6700986 and ITLN1 rs95280 genotype variants among BC patient obesity/IR factors

The presence of obesity, IR or DM, as well as having experienced a pre-diabetic event were associated with mutant homozygous (CC) (for obesity and IR only) and heterozygous (CT) *CD295* rs6700986 genotypes (*P* ≤ 0.05; Table 3). Also, obesity, pre-diabetic event and being diabetic, in addition to IR were associated with BC cases that were *ITLN1* rs952804 mutant heterozygous (CT) (*P* ≤ 0.05; Table 3).

**Table 3.**
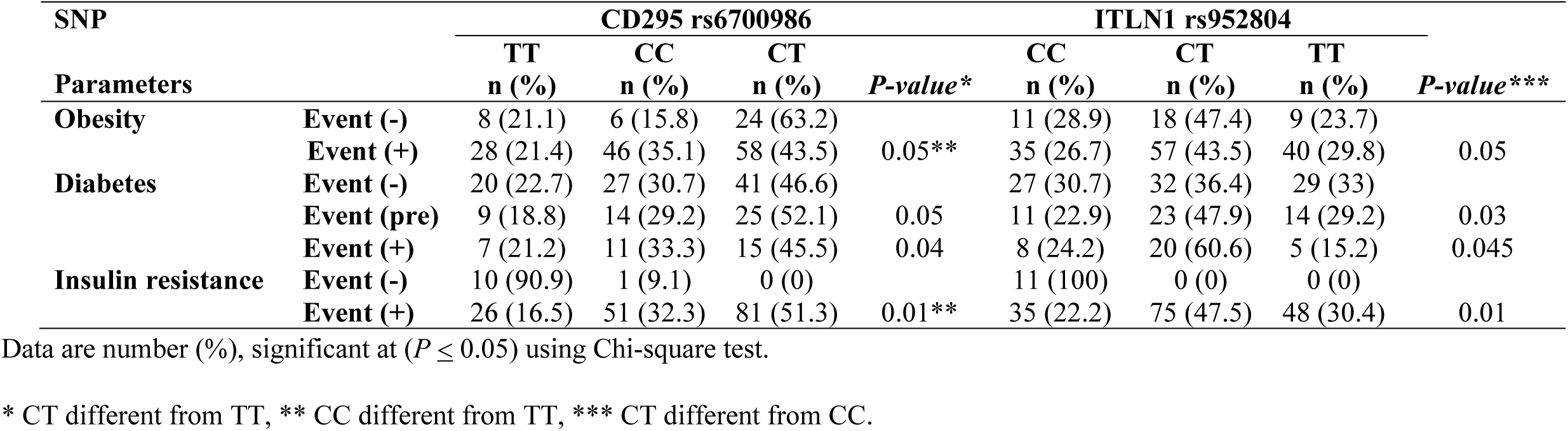
Distribution of genotype variants of CD295 rs6700986 and ITLN1 rs952804 SNPs among obesity/IR factors in breast cancer patients (n=170)

### 3.5. Association between CD295 rs6700986 and ITLN1rs952804 polymorphism and anthropometric parameters of BC patients

Mutant homozygous (CC) and heterozygous (CT) genotypes for *CD295* rs6700986 and *ITLN1* rs952804 mutant homozygous (TT) and heterozygous cases (CT) were significantly (*P* ≤ 0.05) associated with higher age (median 54, 53, respectively) for both SNPs tested (Table 4). This association also occurred for BC patients who had decreased plasma CEA (ng/mL) levels, increased insulin (µIU/mL) levels, IR, or increased plasma leptin (ng/mL) levels (Table 4).

**Table 4.** Association between CD295 rs6700986 and ITLN1rs952804 polymorphisms and anthropometric parameters in breast cancer patients (n=170)

| SNP<br>Parameters | CD295 rs6700986 |  |  |  | ITLN1 rs952804 |  |  |  |
| --- | --- | --- | --- | --- | --- | --- | --- | --- |
|  | TT | CC | CT | *P-value* *** | CC | CT | TT | *P-value* *** |
| Age (years) | 50 (47-64) | 54 (43-75) | 53 (46-75) | 0.05 | 50 (47-75) | 53 (45-75) | 54 (43-75) | 0.001*** |
| CEA (ng/mL) | 4.5 (1-10) | 3.5 (1-10) | 4 (1-10) | 0.029 | 9 (1-10) | 4 (1-10) | 3.8 (1-10) | 0.003 |
| Insulin (μIU/mL) | 7.2 (3.8-16.1) | 13.7 (4.9-17.3) | 10.4 (5.1-15.8) | 0.001 | 8.1 (3.8-13.1) | 10.4 (6-15.8) | 14.4 (9.5-17.3) | 0.001 |
| HOMA-IR | 2.4 (0.9-4.5) | 3.8 (1.2-6.6) | 3.3 (2.5-4.3) | 0.001 | 2.5 (0.9-3.2) | 3.4 (2.7-4.1) | 4 (3.4-6.6) | 0.0001 |
| Leptin (ng/mL) | 15 (8-19) | 16 (11-50) | 15 (8-20) | 0.003** | 15 (8-20) | 15 (8-50) | 16 (11-25) | 0.015** |

Data are median (range), significant at $P \leq 0.05$ using Chi-square test.
\* CT different from TT, \*\* CC different from TT, \*\*\* CT different from CC.
HOMA-IR; homeostasis model assessment-insulin resistance, ITLN1; intelectin-1, CEA; carcinoembryonic antigen, CD295; cluster of differentiation 295.

### 3.6. Relationship between studied SNPs CD295 rs6700986 and ITLN1 rs952804 genotypes and DNA damage parameters

Among DNA damage parameters, only tail DNA (%) and tail moment unit were significantly (*P* ≤ 0.05) associated with *CD295* rs6700986 mutant heterozygous (CT) genotype and *ITLN1* rs952804 mutant homozygous (TT) genotype (Table 5).

**Table 5.** Genotype distribution of CD295 rs6700986 and ITLN1 rs952804 SNPs relative to DNA damage parameters in breast cancer patients (n=170)

| SNPs/<br>DNA damage | CD295 rs6700986 |  |  | *P-value\** | ITLN1 rs952804 |  |  | *P-value\*\** |
| --- | --- | --- | --- | --- | --- | --- | --- | --- |
|  | TT | CC | CT |  | CC | CT | TT |  |
| Tailed (%) | 13 (8-19) | 13 (8-17) | 13 (8-19) |  | 13 (8-19) | 13 (8-19) | 13 (8-17) |  |
| Tail length (µm) | 3.8 (2.7-4.3) | 3.4 (3-4.3) | 3.5 (2.7-4.3) |  | 3.6 (2.7-4.3) | 3.6 (2.7-4.3) | 3.5 (3-4.3) |  |
| Tail DNA (%) | 3.7 (2.5-5) | 4 (2.5-5) | 3.5 (2.5-5) | 0.043 | 4 (2.5-5) | 4 (2.5-5) | 3.4 (2.5-5) | 0.029 |
| Tail moment unit | 12.2 (9.3-17) | 12.2 (10-17.1) | 11.6 (9.3-17) | 0.031 | 12.2 (10-17.1) | 12.2 (10-17.1) | 11 (9.3-16.1) | 0.016 |

Data are median (range), significant at $P \leq 0.05$ using Chi-square test.
\* CT different from TT, \*\* TT different from CC.

## 4. DISCUSSION

The results of our study on a population of BC patients in Egypt demonstrated that the *CD295* gene is highly polymorphic among BC patients. We found that *CD295* polymorphisms were associated with BC cases having late TN classification, as well as FSH levels > 25 mIU/L and HER2/Neu score of +3. Furthermore, BC patients who were obese, diabetic, or pre-diabetic were more likely to carry type I cytokine receptor; leptin receptor (LEP-R) *CD295* polymorphisms. Those patients who had mutant homozygous (CC) or heterozygous (CT) *CD295* rs6700986 genotypes had a higher incidence of tumor clinical stage T II, positive LN involvement, histologic grade III, obesity, pre-diabetes, IR or DM. The CD295 protein is encoded by a gene belonging to the 130-member family of glycoprotein cytokine receptors that are known to enhance gene transcription by activating cytosolic STAT proteins (Vona-Davis and Rose 2007). In addition to functioning in hemoatopoietic pathways that are required for lymphopoiesis, CD295 functions as a receptor for leptin, a hormone that acts on adipocytes to regulate body weight (Wauman et al 2017). Indeed, mutations in the gene that encodes leptin have been related to an increased frequency of obesity (Mahmoudi et al 2015). Moreover, leptin plays a role in cell growth and differentiation, as well as angiogenesis by promoting increased production of signaling molecules such as NO, VEGF, FGF2, and VEGFR2 by endothelial cells (Gonzalez et al 2006). In cancer, leptin is considered to act as both a mitogenic and migration factor in malignant cells (Choi et al 2005). In the context of BC, however, Woo et al (2006) found no significant connection between four polymorphisms in the leptin receptor gene and the risk of BC. They likewise did not observe noteworthy differences in serum leptin levels between BC patients and controls. We also examined the emerging role of *ITLN1* SNPs in BC, and found that rs952804 polymorphisms were related to BC risk factors (grade III, positive LN involvement) as well as to the presence of obesity, IR, DM, and pre-diabetes. These findings are consistent with earlier reports showing that ITLN1 participates in immune defense and insulin-stimulated glucose uptake in human subcutaneous and omental adipocytes (Li et al 2015). In particular, BC cases that were heterozygous for the *ITLN1* rs952804 mutant (CT) were significantly associated with tumor clinical stage T II and positive LN involvement as well as tumor histologic grade III, presence of obesity, pre-diabetic event, DM and IR (*P* ≤ 0.05). Relative to the adiopkine vaspin, ITLN1 levels appear to give be superior indicators of IR among obese patients in the study group by Sperling et al (2016). Obesity is an important health problem, and is positively correlated with the occurrence and mortality of BC (Lorincz and Sukumar 2006, Chu et al 2019). Moreover, obese BC patients are known to have a higher risk of LN metastasis, larger tumors, and higher death rates compared with non-obese patients (Rose and Vona-Davis 2010). These increased risks might be due to elevated estrogen levels that occur from enhanced aromatization in adipose tissue and increases in the levels of mitogenic agents such as insulin and/or IGF associated with obesity-related metabolic syndromes (DM and BC) (Erbay et al 2016). In addition to increased estrogen levels, increases in insulin levels may also contribute to the development of BC by enhancing insulin-like growth factor-I (IGF-1) receptor-mediated release of VEGF from breast tissues (Erbay et al 2016), which could couple with effects associated with increased leptin levels (Belfiore 2007). Moreover, the adverse influence of DM on the prognosis of cancer patients with malignancies is likely associated with effects of insulin on the tyrosine kinase growth receptor pathway (Belfiore 2007). Insulin, IGF-I, and hybrid IGF-I/insulin receptors, are all overexpressed in BC cells (DeCensi and Gennari 2011). As such, activation of these receptors could up-regulate expression of insulin receptor substrate 2 that in turn activates downstream MAPK and phosphatidylinositol 3-kinase-Akt pathways (Belfiore 2007). Indeed, *in vitro* studies demonstrated that ITLN1 increases insulin signal transduction by activating protein kinase Akt/protein kinase B signaling and enhancing insulin-stimulated glucose transport in isolated human adipocytes. Increased BC risk related to hyperinsulinemia could be attributed to a synergistic interaction between elevated free estrogen concentrations and aberrant insulin signaling. Furthermore, the ability of ITLN1 to activate AKT, a key survival factor, could be a mechanism for ITLN1-mediated cell proliferation in BC (Bahadori et al 2014). This effect, together with CD295 stimulation of gene transcription associated with activation of cytosolic JAK-STAT proteins, MAPK, PI3K, and AMPK (Mahmoudi et al 2015), may contribute to the increased frequency of *ITLN1* and *CD295* polymorphisms seen in this study. It should be noted that *ITLN1* rs952804 mutant heterozygous (CT), specifically the T allele, was related to BC risk, DNA damage and obesity, DM and IR. In addition, there was no statistically significant difference between our observations and that predicted by the Hardy-Weinberg principle for *ITLN1*, suggesting that the population was in Hardy-Weinberg equilibrium (HWE) (data not shown). Thus, the null hypothesis is *not* rejected, and we presume that the population is indeed in HWE, although additional study is needed to contrast the findings for this locus with that of another study population. There is currently no BC screening program in Egypt and in the absence of population-based-cancer registries in developing nations, including Egypt, patients with early pre-clinical illness may have been missed in our study. It is possible that polymorphisms of *CD295* or potentially *ITLN1* could contribute to variations in BC occurrence. Nonetheless, in this study SNPs in *CD295* and *ITLN1* were related to oxidative stress/DNA damage in postmenopausal Egyptian female BC patients.

## 5. Conclusions

This study showed an increased risk of BC in women carrying the C allele of the *CD295* rs6700986 polymorphism and the T allele of the *ITLN1* rs952804 polymorphism. Our results suggest that these SNPs could be considered as BC-related risk factors.

### RECOMMENDATIONS

*ITLN1* and *CD295* polymorphism testing could be used to assess BC susceptibility in either obese or insulin resistant, pre-diabetic patients. We therefore suggest that this high-risk BC population be screened for these SNPs.

## Data Availability

Available

## Author contribution

Authors contributed equally to all aspects of this study, including data collection and analysis and drafting of the manuscript.

